# An Investigative study of methods for Retinal Image Registration

**DOI:** 10.64898/2025.12.01.25341352

**Authors:** Thenuka Dharmaseelan, Neelabh Sinha, Samyyia Ashraf, Kimia Daneshvar, Yiu Wai Chan, Nikolas Pontikos

## Abstract

**Aim:** This study aims to compare three deep learning-based retinal image registration methods RetinaRegNet, EyeLiner, and GeoFormer on the FIRE dataset to determine which approach provides optimal registration accuracy and computational efficiency across varying image overlap conditions (Classes S, A, and P) using mean landmark error as the primary outcome measure.

**Methods:** The three pipelines were evaluated under consistent conditions. RetinaRegNet incorporates diffusion features, dual keypoint sampling (SIFT and random), two stage outlier removal, and a multilevel registration hierarchy progressing from homography to polynomial transforms. EyeLiner integrates anatomical segmentation with SuperPoint feature extraction, LightGlue matching, and thin-plate spline warping. GeoFormer builds on LoFTR through cross-attention mechanisms and RANSAC-based refinement. Registration performance was quantified using mean landmark error (MLE).

**Results:** Across all 134 FIRE image pairs, RetinaRegNet achieved the lowest overall MLE (3.12 pixels), outperforming EyeLiner (3.66 pixels) and GeoFormer (6.06 pixels). Class-specific analysis showed that RetinaRegNet delivered the highest accuracy in Class S images (1.70 pixels), competitive performance in Class A (5.24 pixels), and the strongest results in the most challenging Class P cases (4.57 pixels). GeoFormer demonstrated the shortest processing time at 0.32 seconds per image pair, compared with 4.92 seconds for EyeLiner and 31.23 seconds for RetinaRegNet. In Class P, RetinaRegNet achieved a 59.2% improvement in accuracy relative to GeoFormer (4.57 vs 11.20 pixels).

**Discussion:** Overall, the evaluation reveals a clear trade-off between registration precision and computational speed. RetinaRegNet achieves the lowest MLE for complex clinical cases despite higher computational cost, EyeLiner balances precision and speed for routine use, while GeoFormer prioritizes rapid throughput where processing speed is critical.

## Introduction

Retinal image registration represents a cornerstone of modern ophthalmic research and clinical care, enabling precise alignment of images across different time points to track disease progression and evaluate treatment efficacy [1, 2]. The human retina provides a unique window into systemic health, with retinal diseases such as age-related macular degeneration (AMD), diabetic retinopathy, and glaucoma affecting millions globally and creating growing healthcare burdens as populations age [2–5]. Retinal imaging techniques including colour fundus photography (CFP), fundus autofluorescence (FAF), infrared reflectance (IR), fluorescein angiography (FA), and scanning laser ophthalmoscopy (SLO) capture complementary anatomical and functional information, essential for comprehensive disease assessment. However, effective utilisation of retinal imaging data depends on robust registration frameworks capable of accurately aligning images despite substantial variations in resolution, contrast, illumination, and spectral characteristics across imaging modalities.

The advent of deep learning precipitated a paradigm shift, initially utilising Convolutional Neural Networks (CNNs) to predict alignment directly or via coarse-to-fine strategies [6–8]. As the field matured, research pivoted toward addressing specific limitations of early CNNs. Style-transfer frameworks were introduced to unify multimodal representations, enabling robust vessel segmentation without pixel-wise annotations [9, 10]. To eliminate the dependency on ground-truth transformations, unsupervised deformable networks employing Spatial Transformer architectures were developed [11], while multi-scale frameworks incorporating edge similarity losses addressed optical distortions in fundus-OCT alignment [12].

Despite these sophisticated advances, the demand for greater robustness and interpretability led to the divergence of three specialized architectural families. First, anatomically guided models began leveraging structural priors; early work explored structure-driven regression [13], while keypoint-based methods focused on explicit landmark matching [14, 15]. Second, transformer-based architectures and detector-free frameworks were adopted to capture long-range spatial dependencies via attention mechanisms [16, 17]. Finally, generative diffusion models, built on foundational probabilistic synthesis research, emerged to synthesize robust, illumination-invariant feature representations [18, 19].

Yet, a clear consensus on the optimal balance between computational efficiency and registration accuracy across these families remains elusive. To address this gap, we conduct a focused comparative analysis of three state-of-the-art frameworks on the FIRE dataset: **RetinaRegNet**, a zero-shot diffusion-based model leveraging hierarchical outlier suppression [20]; **EyeLiner**, an anatomically guided pipeline combining vessel landmark extraction with transformer matching [21]; and **GeoFormer**, a geometry-aware transformer incorporating LoFTR-style correspondence detection with RANSAC filtering for spatial consistency [22]. By evaluating these methods under varying degrees of overlap and anatomical distortion, this study provides a systematic assessment of their robustness, efficiency, and clinical suitability for large-scale retinal image analysis.

### Methodology

#### Dataset Description

The FIRE (Fundus Image Registration) dataset is widely recognized as a benchmark for evaluating retinal image registration algorithms. It comprises 134 colour fundus image pairs derived from 129 individual images, each captured at a resolution of 2912 × 2912 pixels with a 45° field of view using a NIDEK AFC-210 fundus camera [23]. To enable systematic performance assessment, the dataset is divided into three difficulty categories based on field-of-view overlap and anatomical variation: **Class S (71 pairs)** represents easy cases with substantial image overlap (>75%); **Class A (14 pairs)** includes moderate cases exhibiting noticeable anatomical changes (see Figure 1) between images; and **Class P (49 pairs)** comprises the most challenging cases with limited overlap (<75%).Each image pair includes 10 manually annotated ground-truth landmarks, typically located at vessel bifurcations and crossings, which provide a reference standard for quantitative evaluation of registration accuracy. A visual overview of representative image pairs from each FIRE class is shown in **Figure 2**.

**Figure 1.**
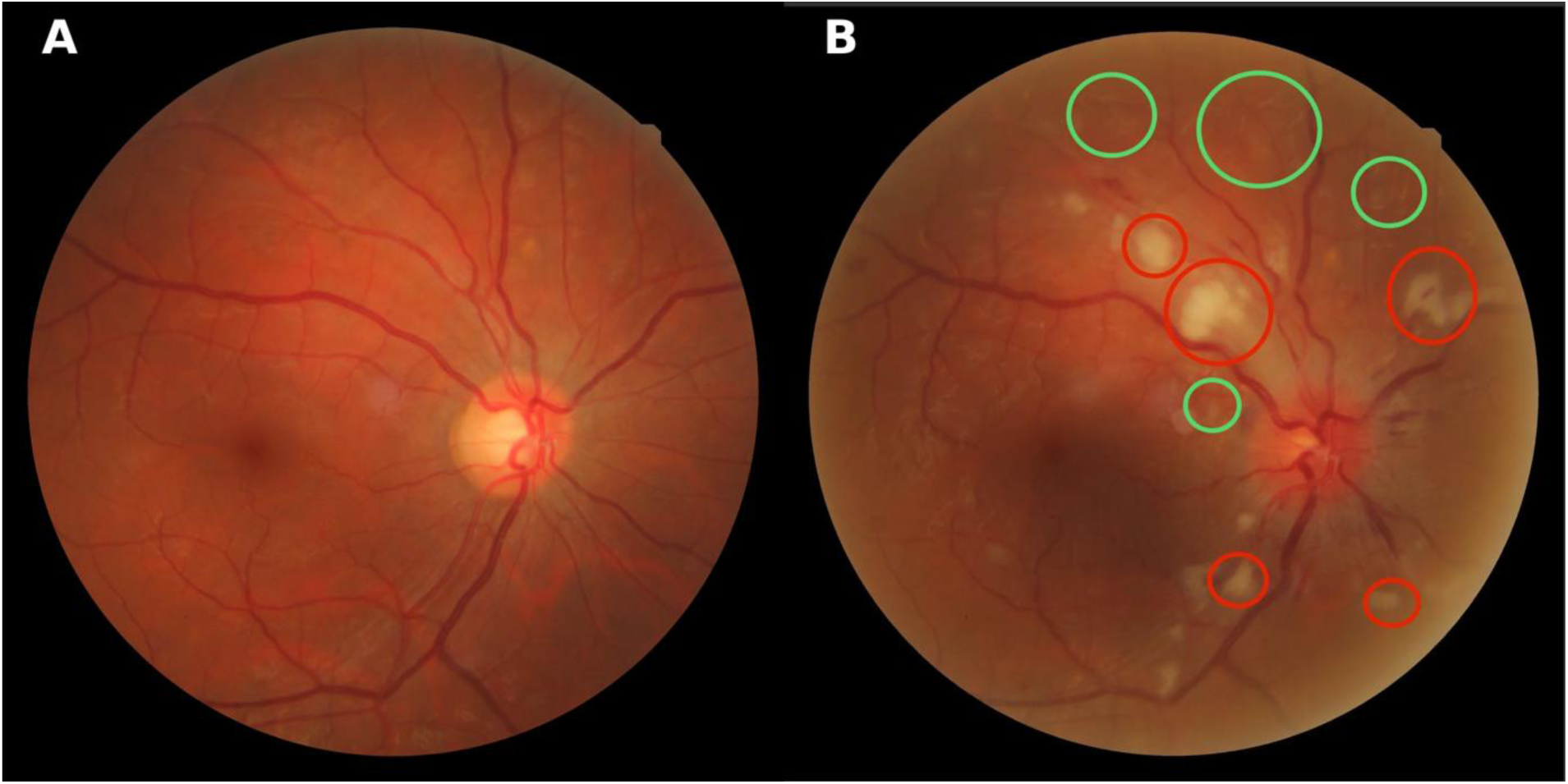
Class A anatomical changes on the FIRE dataset. A: Reference retinal image showing normal fundus appearance; B: Test image with pathological features highlighted. Red circles indicate hard exudates; green circles denote cotton wool spots. Prominent optic disc swelling is observed in the test image.

**Figure 2.**
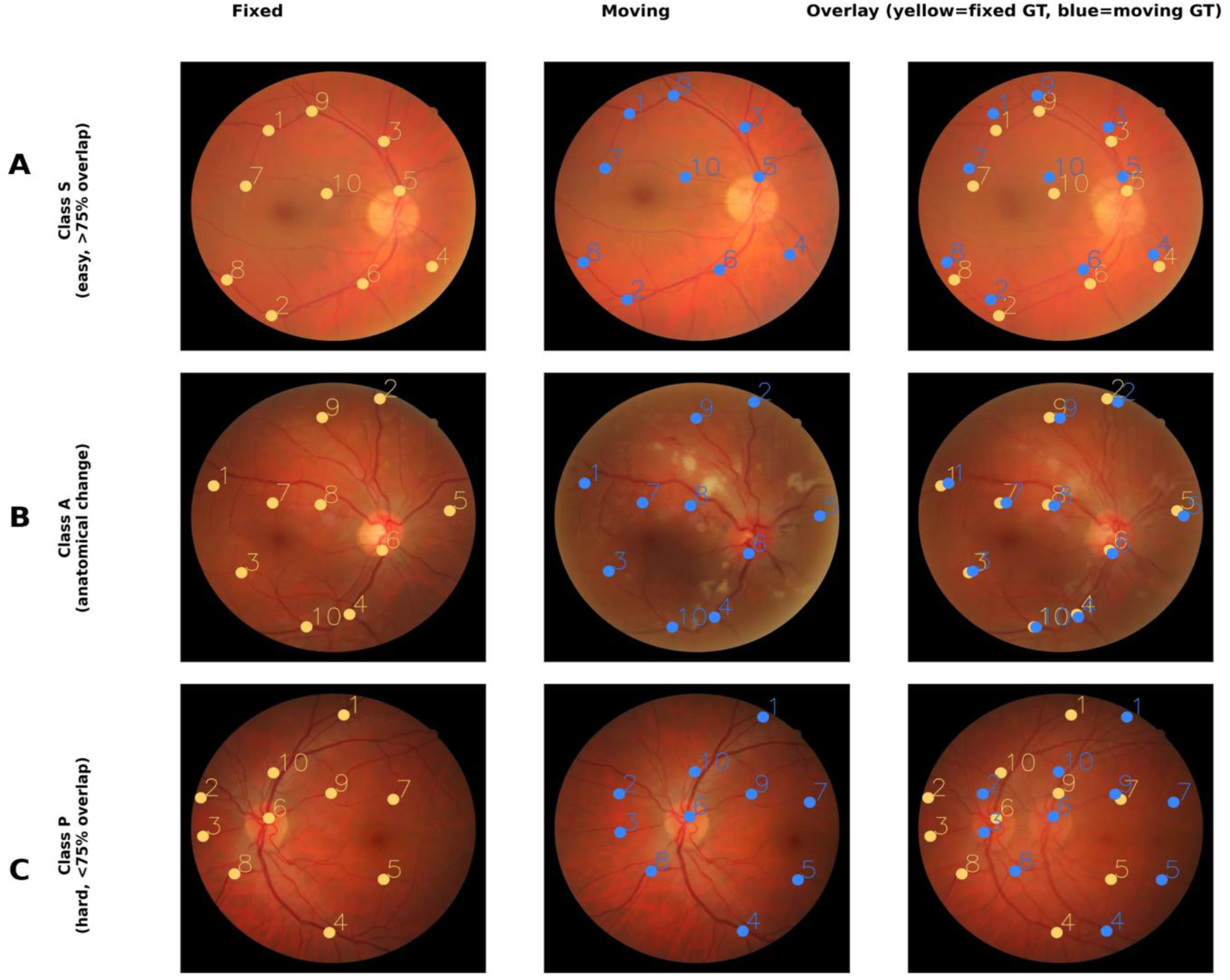
Pre-registration visual summary on the FIRE dataset. A: Class S (*n*=71, easy, >75% overlap); B: Class A (*n*=14, anatomical change); C: Class P (*n*=49, hard, <75% overlap). Columns show the fixed (reference) image, moving (test) image, and their unregistered overlay. Ground-truth landmarks are shown as yellow (fixed) and blue (moving) points. All images are 2912×2912 px, 45◦ FOV, captured with a Nidek AFC-210 fundus camera.

#### RetinaRegNet: Novel Zero Shot Diffusion Based Registration Framework

RetinaRegNet is a zero-shot framework designed to register pairs of retinal images by identifying correspondences within semantic diffusion features and subsequently warping one image onto the other. The process follows a broadly three-stage pipeline encompassing **feature extraction, correspondence refinement, and hierarchical transformation** (**Figure 3**).

**Figure 3.**
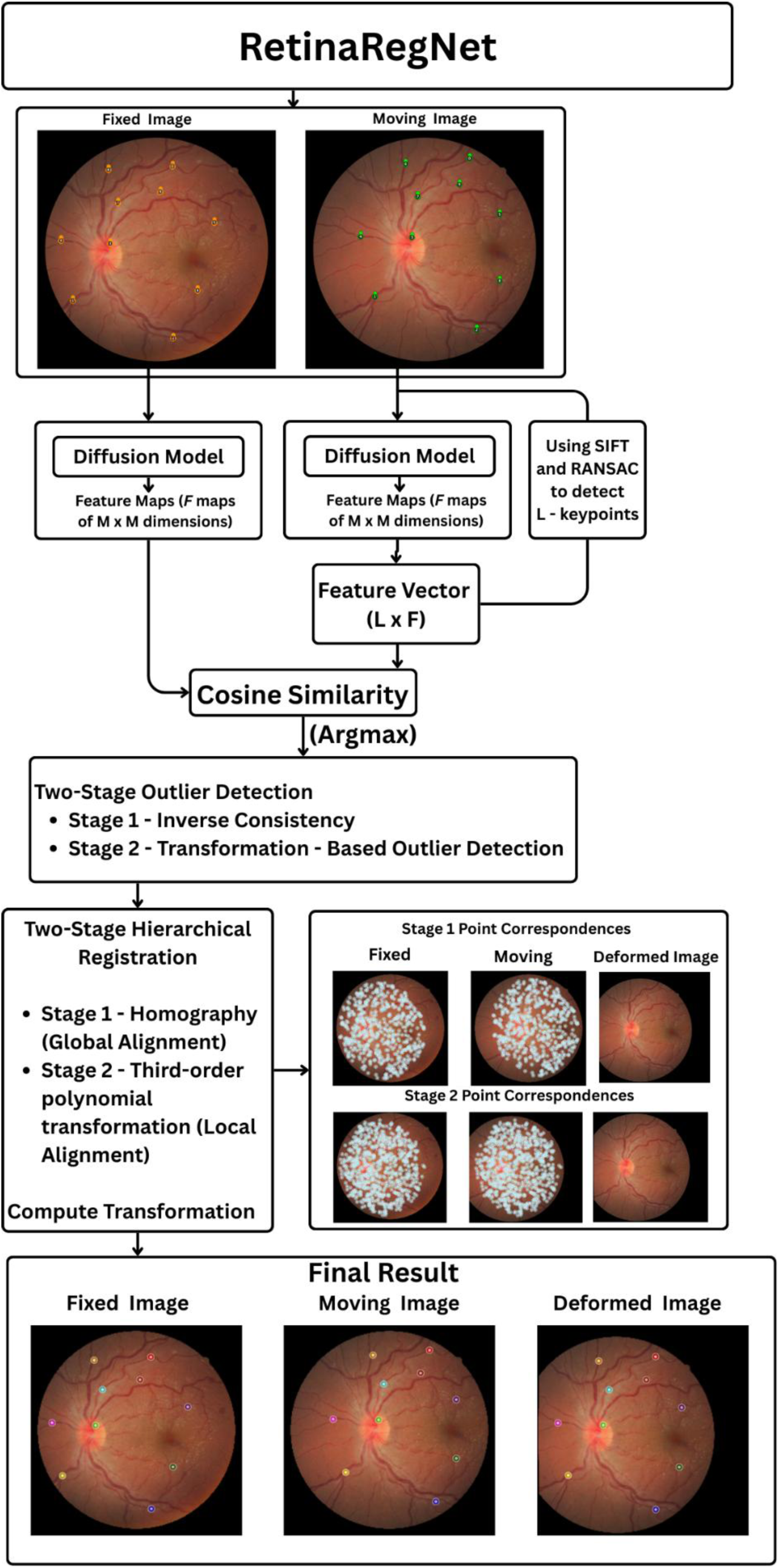
Overview of the RetinaRegNet registration pipeline. Moving and fixed fundus images are processed through diffusion models to extract feature maps. SIFT and RANSAC detect keypoints, and cosine similarity establishes correspondences. Two-stage outlier detection filters unreliable matches. Two-stage hierarchical registration applies homography for global alignment and third-order polynomial transformation for local alignment, producing the final registered output.

First, rich feature maps are extracted from both the fixed and moving images using a **pretrained Stable Diffusion** model. Each image is passed through the model at a low noise step, producing intermediate feature tensors that capture both vessel patterns and broader retinal anatomy while remaining robust to variations in illumination and contrast.

With these features in hand, the method selects a balanced set of control points, approximately half from SIFT (to capture textured vessel regions) and half sampled uniformly at random (to cover smooth areas), ensuring that correspondence estimation is not biased toward densely textured regions. It then computes correspondences by **cosine similarity** in feature space so that a point *p* in the fixed image is paired with the most similar location *q* in the moving image. This approach provides invariance to photometric variations and tolerates moderate geometric drift.

Matched pairs are then refined through a two-step filtering process that eliminates outliers. First, a forward–backward **(inverse consistency)** check ensures that correspondences agree in both directions; mismatches that fail this test are discarded. Second, a **geometric filtering** step removes any pairs that deviate significantly from a globally consistent transformation. This combination of semantic matching and hierarchical outlier rejection produces a dense but reliable correspondence field that can support both global and local alignment.

Finally, RetinaRegNet performs a coarse-to-fine warp in two stages. Stage 1 estimates a **global homography** to absorb overall eye or camera motion, followed by Stage 2, a smooth local polynomial warp to correct small residual deformations. In practice, this combination of semantic feature matching, rigorous yet efficient filtering, and sequential global-to-local warping achieves high registration accuracy, particularly in low-overlap scenarios. The diffusion features enable recognition of vascular and anatomical patterns even where traditional pixel-based methods fail, while the two-stage transformation mitigates overfitting and preserves anatomical coherence.

#### EyeLiner

EyeLiner replicates clinical assessment patterns through modular anatomical structure analysis. The pipeline can be understood as a four-stage process that algorithmically mimics how clinicians track pathological changes relative to stable anatomical landmarks. An overview of the EyeLiner pipeline is shown in **Figure 4**.

**Figure 4.**
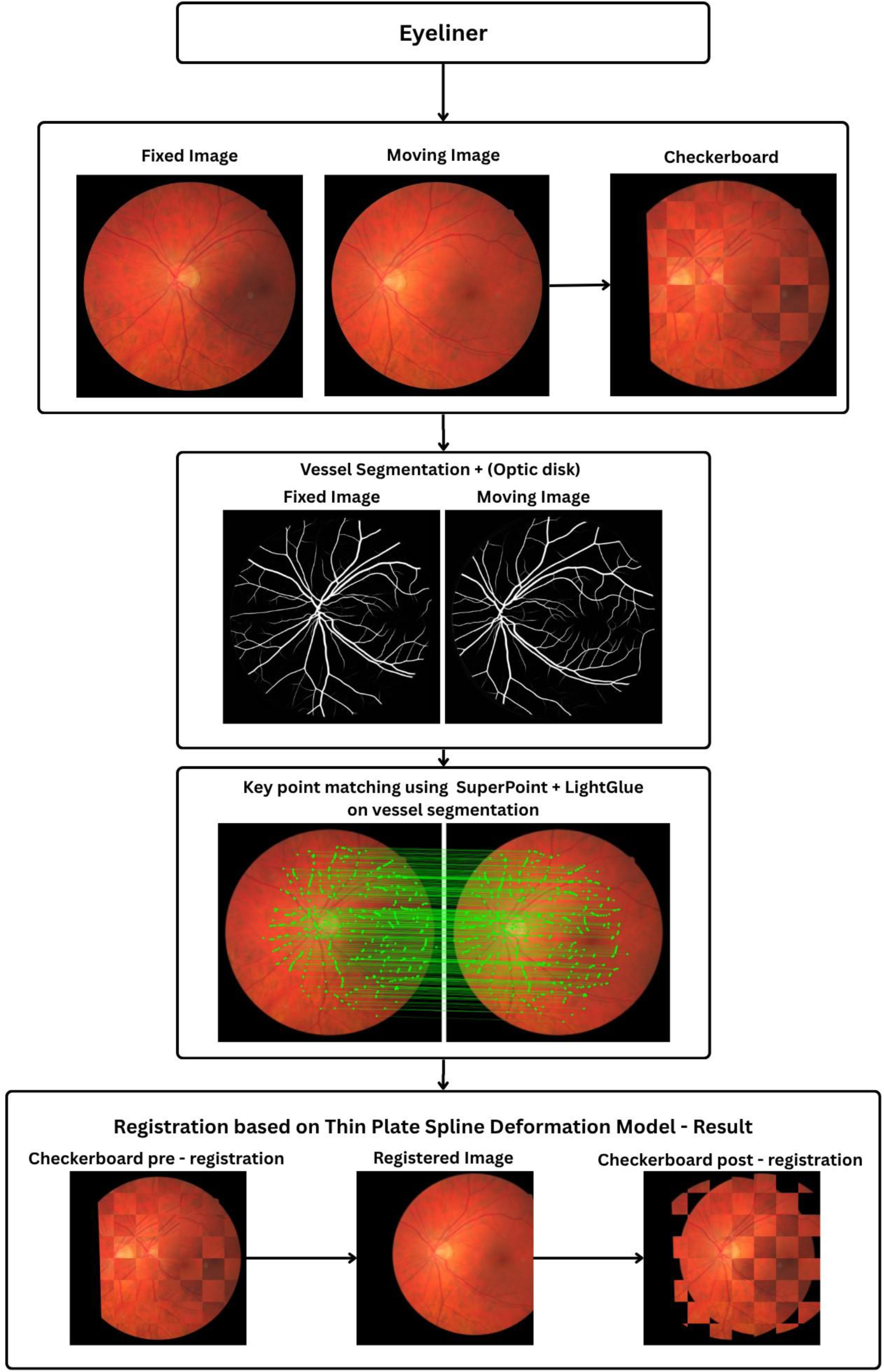
Overview of the EyeLiner registration pipeline. Fixed and moving fundus images are first compared using a checkerboard overlay. Vessel and optic disc regions are segmented to guide correspondence estimation. Keypoints are detected with SuperPoint and matched using LightGlue along vascular structures. A Thin-Plate Spline model then warps the moving image, producing the registered output and improved checkerboard alignment.

The image pairs are first segmented to extract the retinal vasculature and optic disc. The authors employed a standard U-Net architecture trained on established vessel segmentation datasets to generate vascular masks for both the fixed and moving images. For reproducibility, we relied on AutoMorph [24] instead, a publicly available deep learning pipeline for vessel segmentation. Optic disc segmentation is performed using MaskFormer [25], a transformer-based architecture for general-purpose image segmentation. The resulting disc mask can be used to filter out keypoints detected (in the following step) within the optic disc region, as vessels inside the disc are subject to biological motion and thus are less reliable for geometric alignment.

Once the vascular regions are defined, EyeLiner detects and matches distinctive vessel features in a unified process. The SuperPoint [26] network is used to identify salient keypoints along the segmented vessels and to compute descriptor vectors that capture the local appearance of each point. These descriptors are then passed directly to LightGlue [27], a lightweight transformer that performs feature matching. Rather than relying on direct numerical comparison of descriptor values, LightGlue interprets the geometric layout and contextual relationships among vessels in both images, producing more reliable correspondences even when illumination, scale, or focus differ between acquisitions. This combination of SuperPoint and LightGlue therefore establishes anatomically grounded, context-aware correspondences that are robust to the variations that typically challenge conventional intensity-based registration methods.

The final stage of the EyeLiner pipeline performs the actual alignment by warping the moving image to match the fixed reference. Once correspondences are confirmed, a thin-plate spline transformation is used to model the deformation between the two images. This smooth, flexible transformation captures both global displacement and local non-rigid motion in the retinal vasculature while maintaining overall anatomical plausibility. In practice, the combination of anatomically guided segmentation, context-aware matching, and smooth deformation modelling enables EyeLiner to produce clinically interpretable and geometrically stable registrations, offering a strong balance between accuracy, computational efficiency, and biological realism.

#### Geoformer

GeoFormer builds upon the existing LoFTR framework and employs a geometry-aware transformer to learn dense correspondences between retinal images without relying on explicit keypoint detectors. The method can be broadly divided into three stages. A schematic overview of the GeoFormer framework is presented in **Figure 5**.

**Figure 5.**
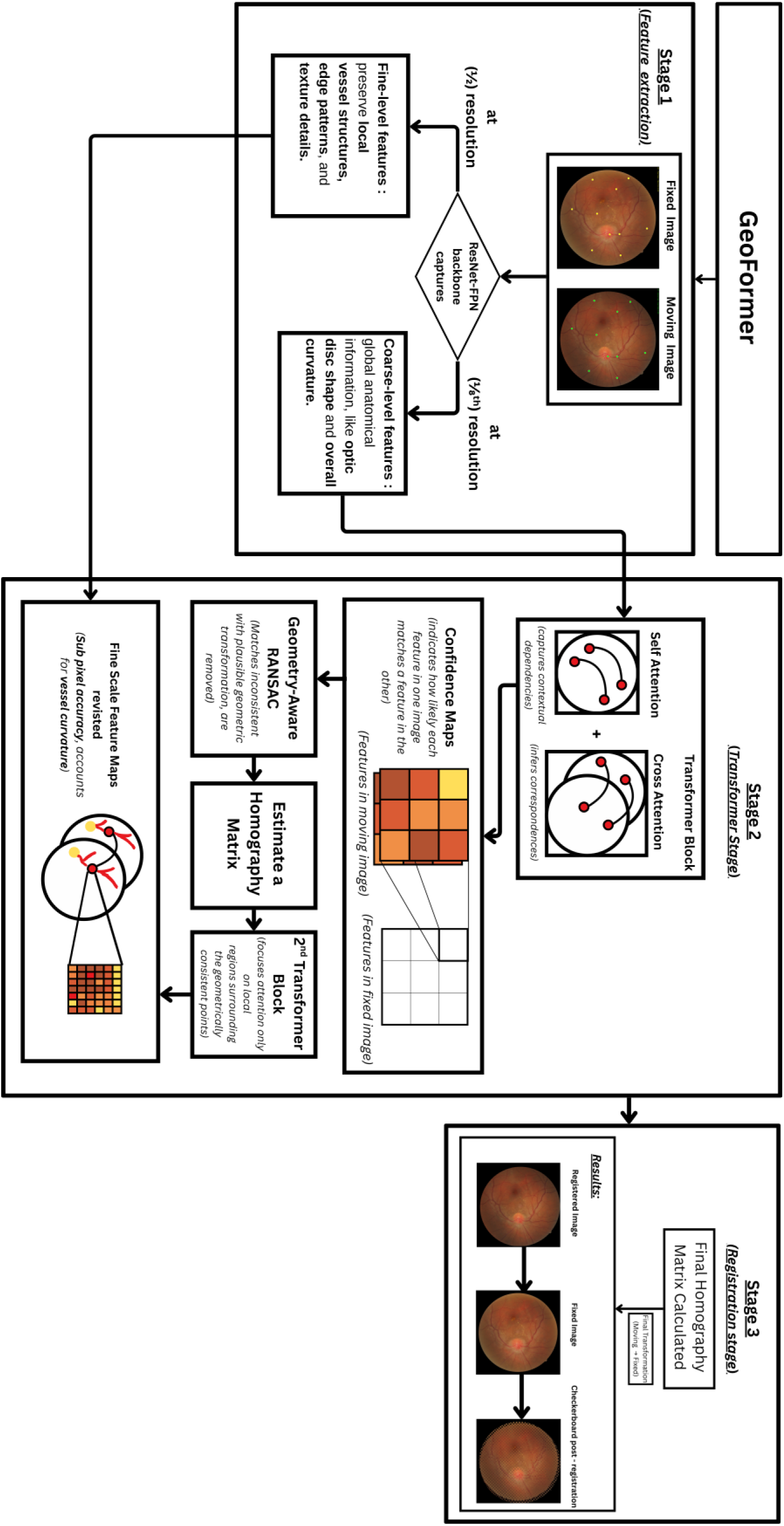
GeoFormer architecture and methodology. Stage 1: ResNet-FPN extracts multi-scale features at 1/2 and 1/8 resolution. Stage 2: Transformer blocks perform self-attention and cross-attention to generate confidence maps, followed by geometry-aware RANSAC and refinement for sub-pixel accuracy. Stage 3: Final homography matrix is computed and applied to produce the registered output.

GeoFormer begins with a ResNet-FPN backbone [28] that extracts multiscale feature maps from the fixed and moving retinal images. The model captures coarse-level features at one-eighth resolution, encoding global anatomical information such as optic disc shape and overall curvature, and fine-level features at one-half resolution, which preserve local vessel structures, edge patterns, and texture details. This multi-level representation ensures that both global context and fine anatomical information are passed to the transformer network.

The extracted features are processed through transformer blocks that perform self-attention within each image to capture contextual dependencies and cross attention between the image pair to infer correspondences. This produces a confidence map that indicates how likely each feature in one image matches a feature in the other. The resulting matches are filtered using a geometry aware RANSAC procedure, which removes correspondences inconsistent with a plausible geometric transformation and estimates an initial homography matrix. These verified matches are then refined through a second transformer stage that focuses attention only on local regions surrounding the geometrically consistent points, improving computational efficiency and accuracy in vessel dense areas. Finally, the model revisits the fine scale feature maps to achieve sub pixel correspondence accuracy, accommodating vessel curvature and illumination variations that coarse features cannot fully capture.

The refined correspondences are used to compute a final homography matrix that maps coordinates from the moving image onto the fixed image. The moving image is then warped according to this transformation, completing the registration process.

#### Evaluation Metrics

Mean Landmark Error (MLE) in pixels for each evaluated method. MLE measures registration accuracy by calculating the average distance between where anatomical landmarks should align versus where the registration method actually places them, with lower values indicating better alignment:

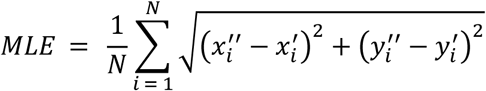

where (*x*′*_i_*,*y_i_*^′^) represent manually annotated ground truth landmarks and (*x*′′*_i_,y_i_*′′) are transformed coordinates.

#### Experimental Setup

**All three registration pipelines were reproduced and evaluated under a unified computational environment to ensure consistency and comparability across methods.** All experiments were executed on a workstation equipped with an **NVIDIA GeForce RTX 3090 GPU (24 GB VRAM)** running **CUDA 12.6**. The implementations used **Python 3.11.13**, **PyTorch 2.2.2**, and **torchvision 0.17.2** as the primary deep learning frameworks. For methods requiring vessel segmentation (EyeLiner), AutoMorph was used for reproducibility. **All runtime and accuracy measurements reported in Results section reflect executions under this standardised environment.**

## Results

### Performance Comparison on FIRE Dataset

Table 1 presents the Mean Landmark Error (MLE) in pixels for each evaluated method on the FIRE dataset. The metrics are reported for the full dataset as well as across three difficulty-based categories: Class S (easy cases with image overlap *>* 75%), Class A (moderate cases with anatomical changes), and Class P (hard cases with image overlap *<* 75%). This breakdown provides insight into method performance under varying levels of image complexity.

**Table 1.** Performance Comparison on FIRE Dataset using Mean Landmark Error (MLE).

| Method | MLE<br>(*n*=134) | Class S MLE<br>(*n*=71) | Class A MLE<br>(*n*=14) | Class P MLE<br>(*n*=49) |
| --- | --- | --- | --- | --- |
| RetinaRegNet | 3.12±2.43 | 1.70±0.54 | 5.24±2.64 | 4.57±2.80 |
| EyeLiner | 3.81±3.13 | 1.80±0.40 | 4.87±3.05 | 6.01±3.75 |
| GeoFormer | 6.06±4.86 | 2.42±0.77 | 6.55±4.71 | 11.20±3.25 |

### Overall Performance Analysis

As shown in Table 1 and Figure 6, RetinaRegNet achieved the lowest overall Mean Landmark Error (MLE) of 3.12 ± 2.43 pixels, demonstrating superior registration accuracy across the FIRE dataset. A Friedman test confirmed a significant overall difference between the three methods (χ²(2)=134.37, *P*<2×10⁻¹⁶), and post-hoc Wilcoxon signed-rank tests with Bonferroni correction showed that RetinaRegNet achieved significantly lower MLE than both EyeLiner and GeoFormer, while EyeLiner significantly outperformed GeoFormer.

**Figure 6.**
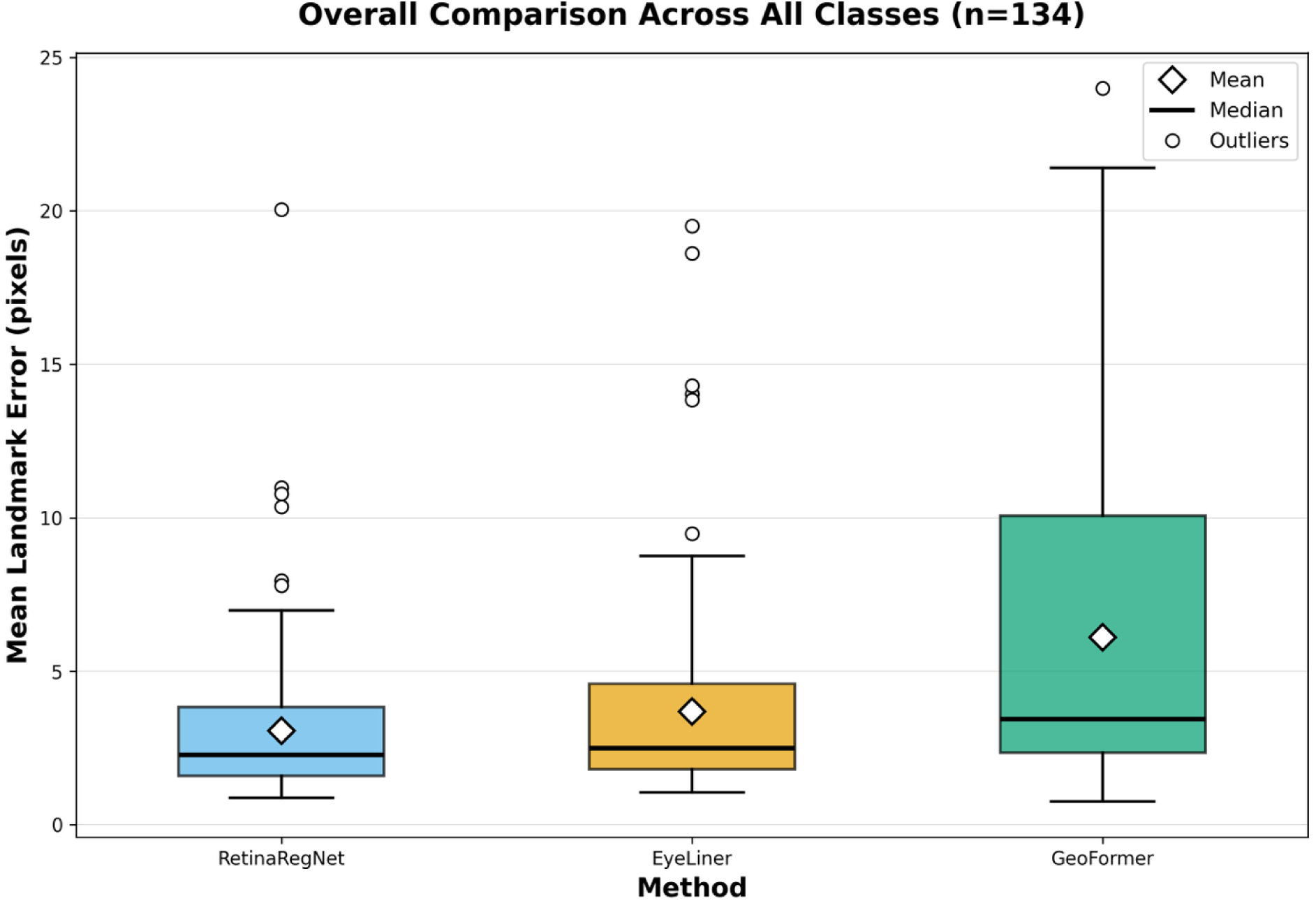
Overall comparison of registration methods across all classes on the FIRE dataset (*n*=134). Box plots show the distribution of Mean Landmark Error (MLE) in pixels for RetinaRegNet, EyeLiner, and GeoFormer methods.

EyeLiner followed with an overall MLE of 3.81 ± 3.13 pixels, which was significantly higher than RetinaRegNet (*P*<1×10⁻⁴). GeoFormer exhibited the highest error at 6.06 ± 4.86 pixels, significantly worse than both RetinaRegNet and EyeLiner (all *P*<1×10⁻¹⁸), indicating reduced registration accuracy.

The box plot analysis in Figure 6 reveals important distributional characteristics of each method’s performance. RetinaRegNet demonstrates the most compact error distribution with a median of approximately 2.0 pixels and relatively few outliers, suggesting consistent performance across the majority of image pairs. EyeLiner shows a slightly wider distribution with a median around 2.5 pixels and a comparable outlier pattern. In contrast, GeoFormer exhibits substantially higher variability, with a median of approximately 4 pixels and a significantly larger interquartile range, indicating less stable performance across different registration scenarios.

These results reveal meaningful differences in both accuracy and reliability among the evaluated methods. While all models demonstrate the capability to perform landmark-based registration, the performance gaps highlight the varying capacities of the underlying architectures to model geometric transformations and generalize across diverse retinal images in the FIRE dataset. The lower mean and standard deviation of RetinaRegNet, combined with its compact error distribution, suggest superior robustness to variations in image quality, pathological features, and geometric complexity.

### Class Specific Performance Analysis

- **Class S Performance:** Class S, representing the largest subset with 71 samples, proves to be the least challenging for all methods Figure 7A. RetinaRegNet obtains the best performance in this class with an MLE of 1.70 pixels, outperforming EyeLiner (1.80 pixels) and GeoFormer (2.42 pixels) by 6% and 42%, respectively. The consistently low error rates across all methods indicate that Class S images possess favorable characteristics for vessel landmark localization. These differences were statistically significant (Friedman *P*=2×10⁻¹⁴; all pairwise comparisons were statistically significant after Bonferroni correction (all adjusted *P* ≤ 8×10⁻⁴), indicating that RetinaRegNet provides consistently lower landmark error than EyeLiner and GeoFormer in this class.
- **Class A Performance:** Class A, representing moderate cases with anatomical change (14 samples), shows intermediate difficulty Figure 7B. EyeLiner attains the lowest mean MLE in this class (4.87 pixels), followed closely by RetinaRegNet at 5.24 pixels; however, this difference is not statistically significant (adjusted *P* = 0.776). GeoFormer shows higher error at 6.55 pixels and performs significantly worse than EyeLiner (adjusted *P* = 0.012). These results indicate that performance differences between the methods are less pronounced in this subset, reflecting the moderate difficulty of Class A cases.
- **Class P Performance:** Performance degradation occurs across all methods on Class P samples (49 samples), the most challenging category Figure 7C. RetinaRegNet maintains relatively strong performance with an MLE of 4.57 pixels, while EyeLiner records 6.01 pixels. GeoFormer experiences substantial difficulty on this class, with its error increasing to 11.20 pixels a 145% increase compared to RetinaRegNet. This pronounced performance gap reveals that GeoFormer is particularly sensitive to the challenging characteristics present in low overlap images. All pairwise differences in Class P were statistically significant after Bonferroni correction, with RetinaRegNet outperforming both EyeLiner (adjusted *P* = 6×10⁻⁴) and GeoFormer (adjusted *P* ≤ 4×10⁻⁹), and EyeLiner also significantly outperforming GeoFormer (adjusted *P* = 4×10⁻⁸).
- **Cross Class Analysis:** Across difficulty classes, RetinaRegNet achieves superior overall performance (3.12 pixels), with the lowest MLE in Classes S and P and mean performance in Class A that is close to EyeLiner. EyeLiner attains the lowest mean MLE in Class A, although its advantage over RetinaRegNet is not statistically significant in this subset, and both methods consistently outperform GeoFormer, which shows particularly large errors in Class P. These findings underscore the importance of class specific evaluation in assessing the robustness and generalization capabilities of retinal registration methods.

### Runtime Analysis

Table 6 presents the runtime performance analysis of the evaluated methods. GeoFormer demonstrates the fastest processing speed at 0.32 seconds per image, offering real-time processing capability. EyeLiner achieves moderate computational efficiency at 4.92 seconds per image. This includes 4.22 seconds for AutoMorph vessel segmentation and 0.69 seconds for the registration process itself, making it approximately 15× slower than GeoFormer. RetinaRegNet exhibits the highest computational cost at 31.23 seconds per image, representing a 98× increase compared to GeoFormer and 6.4× increase compared to EyeLiner.

**Figure 7.**
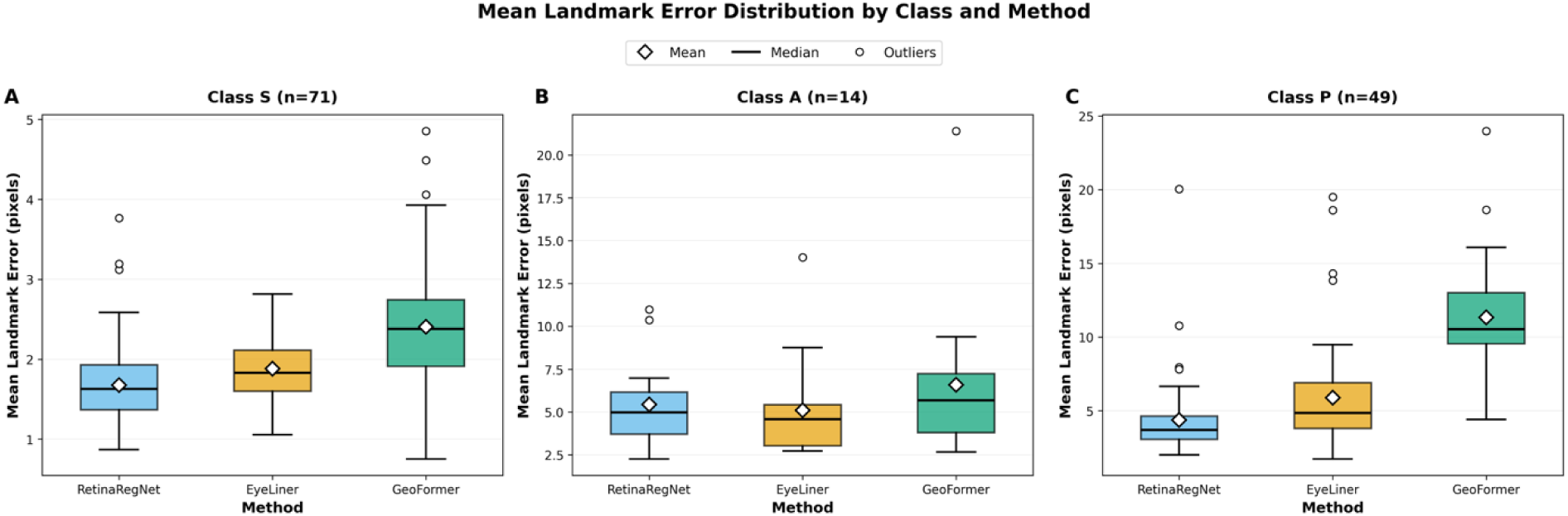
Mean Landmark Error distribution by class and method on the FIRE dataset. Boxplots show the performance comparison of RetinaRegNet, EyeLiner, and GeoFormer across three quality classes. A: Class S (*n*=71); B: Class A (*n*=14); C: Class P (*n*=49). Diamond markers indicate mean values, horizontal lines show medians, boxes represent interquartile ranges (IQR), whiskers extend to 1.5×IQR, and red circles represent outliers. Lower values indicate better registration accuracy.

**Table 2.**
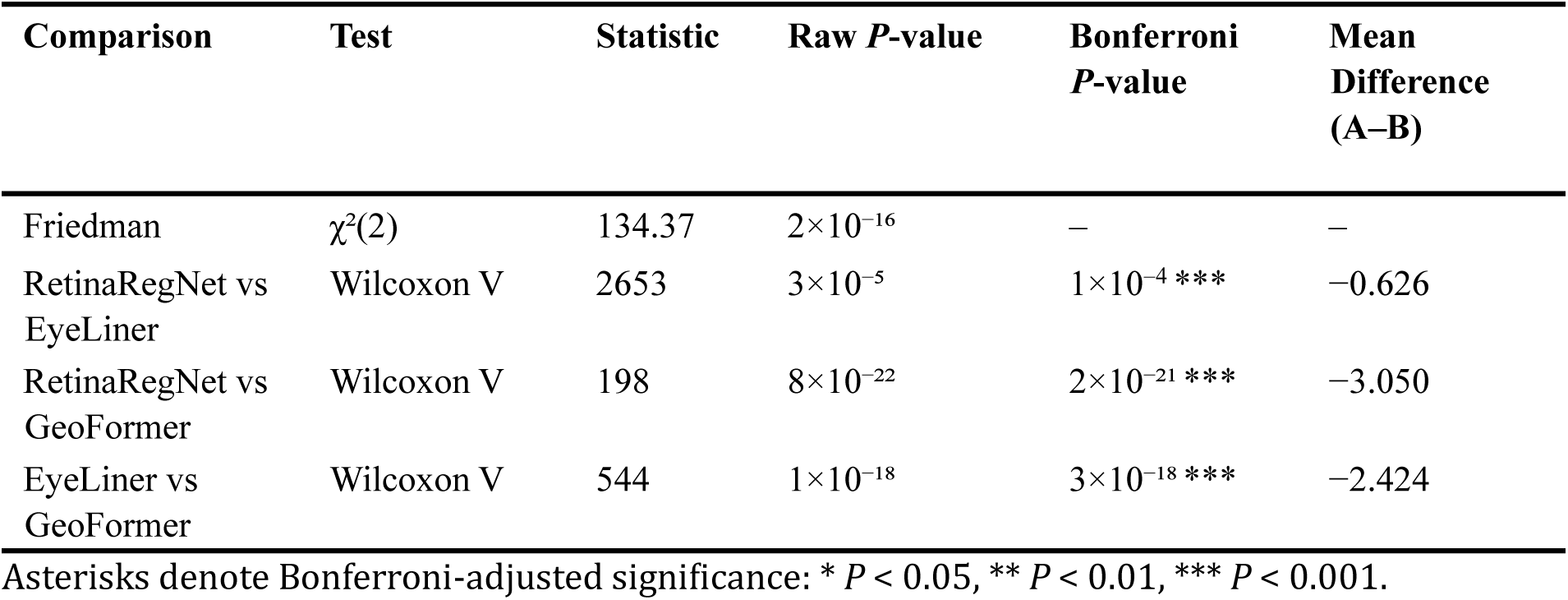
Overall statistical comparison across all methods (*n* = 134)

| Comparison | Test | Statistic | Raw $P$-value | Bonferroni $P$-value | Mean Difference (A-B) |
| --- | --- | --- | --- | --- | --- |
| Friedman | $\chi^2(2)$ | 134.37 | $2 \times 10^{-16}$ | — | — |
| RetinaRegNet vs EyeLiner | Wilcoxon V | 2653 | $3 \times 10^{-5}$ | $1 \times 10^{-4} ***$ | -0.626 |
| RetinaRegNet vs GeoFormer | Wilcoxon V | 198 | $8 \times 10^{-22}$ | $2 \times 10^{-21} ***$ | -3.050 |
| EyeLiner vs GeoFormer | Wilcoxon V | 544 | $1 \times 10^{-18}$ | $3 \times 10^{-18} ***$ | -2.424 |

Asterisks denote Bonferroni-adjusted significance: \* $P < 0.05$ , \*\* $P < 0.01$ , \*\*\* $P < 0.001$ .

**Table 3.** Class S (*n* = 71) statistical comparison of registration methods.

| Comparison | Test | Statistic | Raw $P$-value | Bonferroni $P$-value | Mean Difference (A-B) |
| --- | --- | --- | --- | --- | --- |
| Friedman | $\chi^2(2)$ | 63.577 | $2\times 10^{-14}$ | – | – |
| RetinaRegNet vs EyeLiner | Wilcoxon V | 640 | $2\times 10^{-4}$ | $8\times 10^{-4}$ *** | –0.204 |
| RetinaRegNet vs GeoFormer | Wilcoxon V | 51 | $2\times 10^{-12}$ | $6\times 10^{-12}$ *** | –0.729 |
| EyeLiner vs GeoFormer | Wilcoxon V | 339 | $8\times 10^{-8}$ | $2\times 10^{-7}$ *** | –0.524 |

Asterisks denote Bonferroni-adjusted significance: \* $P < 0.05$ , \*\* $P < 0.01$ , \*\*\* $P < 0.001$ .

**Table 4.**
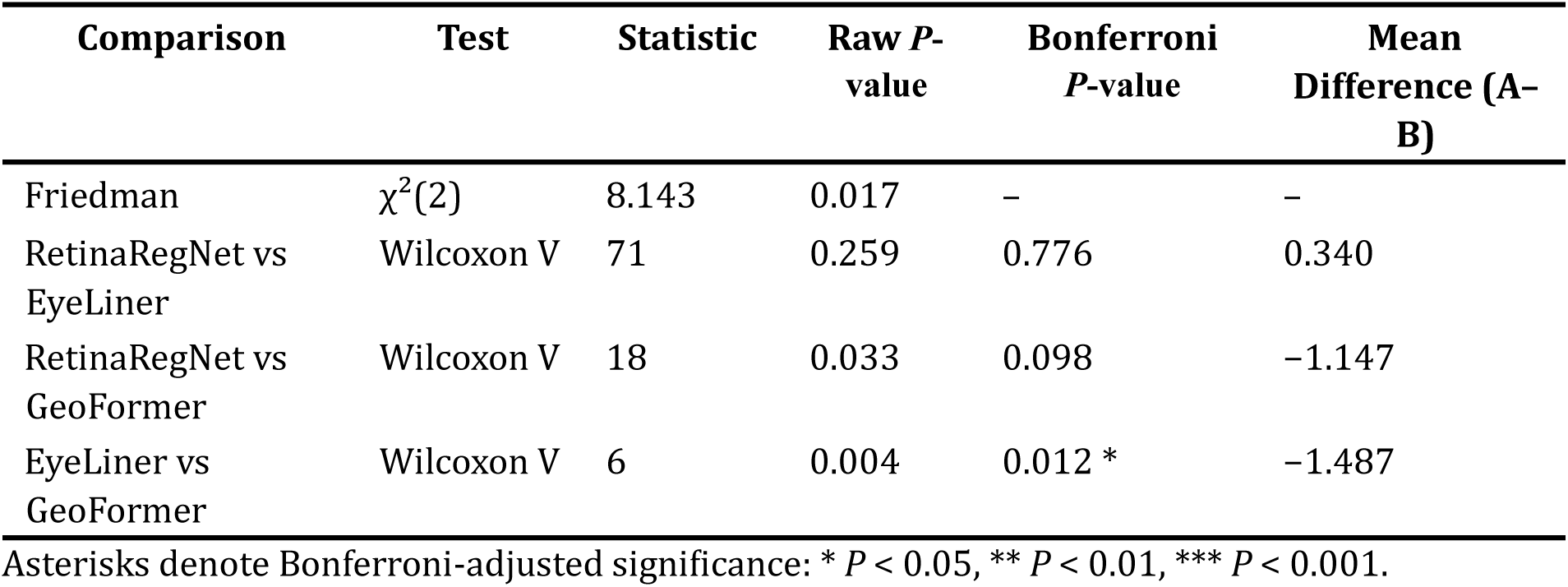
Class A (*n* = 14) statistical comparison of registration methods.

**Table 5.**
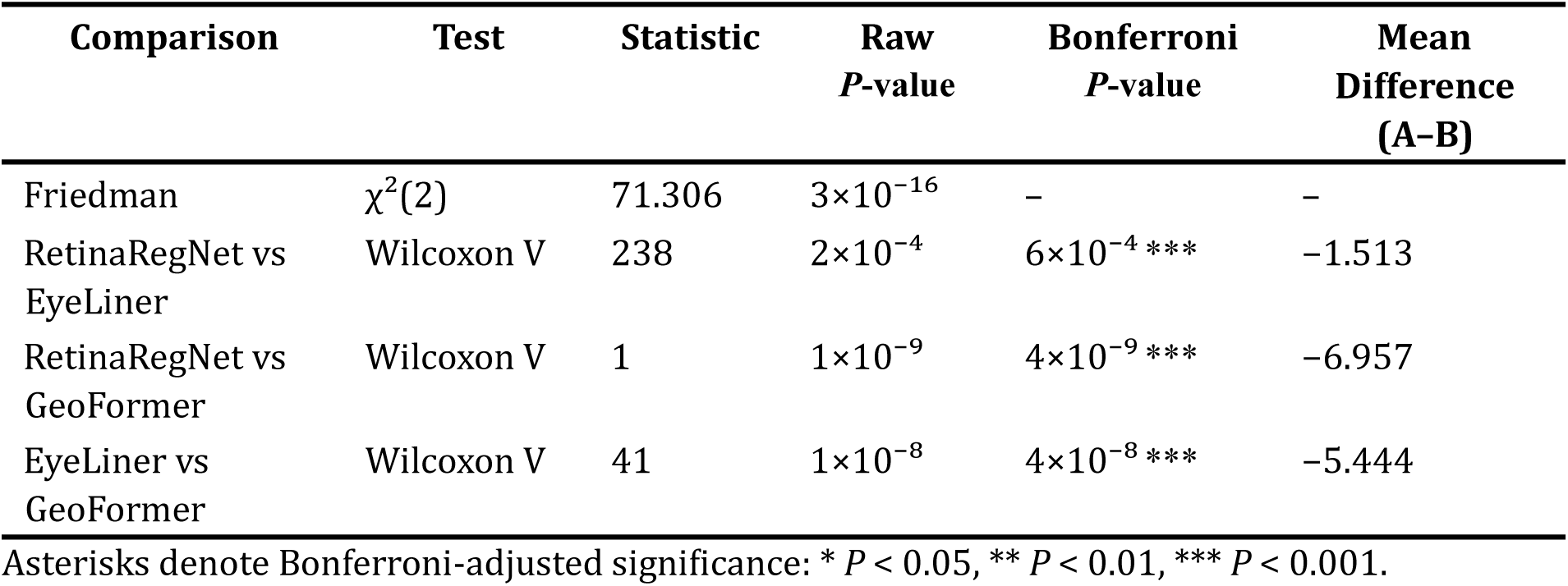
Class P (*n* = 49) statistical comparison of registration methods.

**Table 6.** Runtime Performance Comparison.

| Method | Time per Image<br>(seconds) |
| --- | --- |
| RetinaRegNet | 31.23 |
| EyeLiner | 4.92 |
| GeoFormer | 0.32 |

These computational differences highlight the trade-offs between different architectural approaches. EyeLiner’s reliance on explicit vessel segmentation via AutoMorph adds substantial preprocessing overhead compared to end-to-end approaches. Due to its complex architecture, RetinaRegNet’s multi-stage registration pipeline with iterative refinement steps results in considerably longer processing times. In contrast, GeoFormer’s transformer-based architecture enables direct processing of raw retinal images without intermediate segmentation steps, resulting in significantly faster inference times suitable for clinical deployment.

### Summary of Results

Across the FIRE dataset, the three pipelines demonstrate distinct performance characteristics. RetinaRegNet achieves the lowest overall Mean Landmark Error, indicating strong capability in aligning anatomical landmarks even under challenging imaging conditions. EyeLiner produces competitive accuracy with more stable behaviour across the three difficulty classes. GeoFormer offers very fast processing but displays substantially higher error, particularly in cases with reduced overlap or anatomical distortion. These findings confirm that architectural choices, such as the use of local deformation models and anatomically guided correspondence strategies, play an important role in determining registration accuracy.

### Methodological Analysis

Our results confirm the findings of the previous study [20], showing that incorporating local deformation such as the polynomial deformation in RetinaRegNet and the thin plate spline deformation in EyeLiner in the image warping improves global alignment, as reflected in the outstanding performance in both methods. Inverse consistency in RetinaRegNet helps mitigate inconsistent keypoint matches in regions where the underlying diffusion model exhibits low confidence. However, it becomes less effective when the model is highly confident, which is typically the case for well pretrained models. Consequently, its contribution is limited as EyeLiner is able to achieve comparable performance without imposing strong consistency constraints. Anatomy guided filtering in EyeLiner encourages focus on vascular structure, but can be disadvantageous when vessel morphology changes drastically, leaving large regions without matched keypoints. RetinaRegNet alleviates this overemphasis on edges and high contrast regions through random keypoint sampling, ensuring coverage across nearly the whole image. However, this strategy may mislead the warping algorithm in the presence of anatomical changes, resulting in slightly inferior performance in Class A.

## Discussion

The comparative analysis highlights how the differing design choices of the three methods influence their performance across the FIRE dataset. RetinaRegNet leverages diffusion-based feature maps that provide stable mid-level representations under illumination and contrast changes. While this study does not isolate the specific contribution of diffusion features relative to CNN features, the combination of dense correspondence estimation and its two-stage global-to-local warping appears to support its strong performance, particularly in low-overlap cases. This enables the method to maintain accurate alignment in both easy and difficult pairs and maintains a relative performance advantage in Class P, where reduced overlap increases correspondence ambiguity and demands broader contextual awareness. Its random keypoint sampling supplements vessel-based correspondences by extending coverage across the retina and reducing the likelihood of keypoint sparsity.

EyeLiner performs consistently across classes due to its anatomically guided matching strategy. By focusing correspondence estimation on the vascular network, which is generally stable across acquisitions, the method achieves reliable alignment while limiting sensitivity to illumination differences and local artefacts. Its thin-plate spline transformation captures non-rigid motion effectively. However, vessel segmentation can reduce accuracy in cases where pathology alters vessel morphology, which may explain the slight reduction in performance within Class P.

GeoFormer achieves very rapid inference through its transformer-based correspondence estimation and homography-only warping step. While performing adequately in high-overlap (Class S) scenarios, its reliance on a single global transformation limits its ability to compensate for local structural changes. This constraint is reflected in its higher errors in Class A and Class P, where anatomical variation and reduced shared field of view demand more flexible modelling. The method therefore trades accuracy for speed, making it most suitable for scenarios where throughput is the primary constraint.

Overall, methods incorporating flexible local deformation models achieve superior accuracy. RetinaRegNet and EyeLiner use different yet complementary approaches to model local transformations, and both outperform homography-only registration. The balance between accuracy and computational cost remains a central consideration when selecting an appropriate pipeline for retinal image analysis.

The observed accuracy differences have implications for the integration of retinal registration into clinical workflows. RetinaRegNet’s strong performance in difficult cases supports its use in longitudinal monitoring where reliable alignment is essential for detecting subtle structural change. EyeLiner provides a practical middle ground by delivering competitive accuracy at a lower computational cost and retaining clear anatomical interpretability, making it well suited for routine follow-up assessments and vessel-centric analysis. GeoFormer’s speed makes it appropriate for high-volume contexts such as screening programmes or acquisition quality checks, provided that challenging cases are flagged for secondary processing with a more accurate pipeline. Selecting the appropriate method therefore depends on the specific balance required between precision and computational efficiency.

This study is limited by its reliance on the FIRE dataset, which contains only colour fundus photographs captured with a single 45-degree field-of-view camera at a fixed resolution. Results may differ for other modalities such as autofluorescence, infrared, fluorescein angiography, or widefield imaging. Evaluation was based on ten manually annotated vascular landmarks per pair, which capture alignment accuracy at a limited number of points and may not fully reflect performance across the entire retina, especially near lesions or in peripheral regions. Pixel-based errors were not converted into physical units, which restricts direct clinical interpretation across devices with different magnifications or sensor characteristics.

This study presents a comparative evaluation of three deep learning based retinal registration pipelines using the FIRE dataset. RetinaRegNet achieves the highest accuracy across the full dataset and in the most challenging low-overlap cases. EyeLiner provides reliable performance with moderate computational cost through anatomically guided matching and flexible warping. GeoFormer offers the fastest processing but at the expense of reduced robustness in difficult scenarios. These findings underscore the value of combining global alignment with locally adaptive deformation when precise registration is required and highlight the importance of selecting a registration method that aligns with the clinical or research needs of the intended application. Future work should evaluate these approaches across diverse imaging modalities and optimise high-accuracy models for improved computational efficiency.

## Data Availability

The FIRE (Fundus Image Registration) dataset used in this study is publicly available at https://projects.ics.forth.gr/cvrl/fire/. The source code for RetinaRegNet, EyeLiner, and GeoFormer are available from their respective published repositories. Implementation details and evaluation scripts are available upon reasonable request to the authors.

## Abbreviations

CFP: Colour Fundus Photography
FA: Fluorescein Angiography
FAF: Fundus Autofluorescence
FIRE: Fundus Image Registration (dataset)
FOV: Field of View
IR: Infrared Reflectance
MLE: Mean Landmark Error
SLO: Scanning Laser Ophthalmoscopy
IQR: Interquartile Range
LoFTR: Local Feature Transformer
RANSAC: Random Sample Consensus
SIFT: Scale-Invariant Feature Transform
TPS: Thin-Plate Spline
U-Net: U-shaped Network (architecture)
ICP: Iterative Closest Point
SD: Standard Deviation
API: Application Programming Interface
CUDA: Compute Unified Device Architecture
GPU: Graphics Processing Unit
VRAM: Video Random Access Memory

## Declarations

### Author contributions

**T Dharmaseelan:** Conceptualization, Methodology reproduction, Data curation, Computation and runtime analysis, Results preparation, Writing–original draft, Writing–review & editing, Visualization, Final manuscript preparation.

**N Sinha:** Conceptualization, Literature review, Methodology, Statistical analysis, Data curation, Visualization, Writing–original draft, Writing–review & editing, Final manuscript preparation.

**YW Chan:** Supervision, Conceptualization, Methodology, Validation, Writing–review & editing. Final manuscript preparation.

**S Ashraf:** Supervision, Writing–review & editing, Manuscript editing, Final draft review.

**K Daneshvar:** Literature review, Writing–original draft (clinical implications and limitations), Writing–review & editing.

**N Pontikos:** Supervision, Project administration, Validation, Writing–review & editing, Final approval of the manuscript.

**All authors read and approved the submitted version.**

## Ethical Approval

Not Applicable

## Consent to Participate

Not Applicable

## Availability of data and materials

The **FIRE dataset** analysed during this study is publicly available at: https://projects.ics.forth.gr/cvrl/fire/. No additional datasets were generated or analysed. Code is available upon reasonable request.

## Supplementary

**Supplementary Figure 1.**
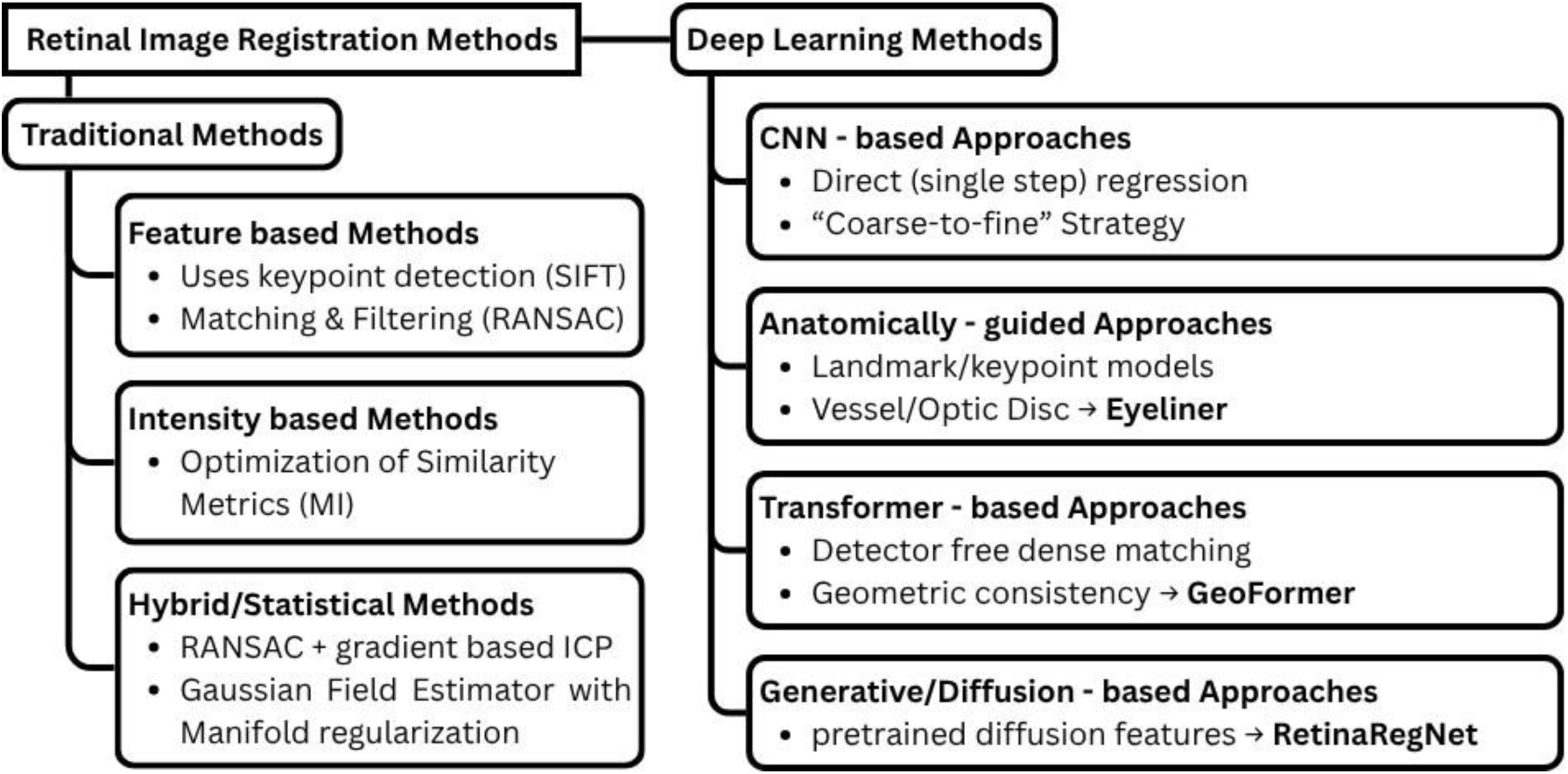
Taxonomy of retinal image registration methods. The field can be broadly divided into two main families: **Traditional methods** and **Deep Learning methods**

**Supplementary Table 1.**
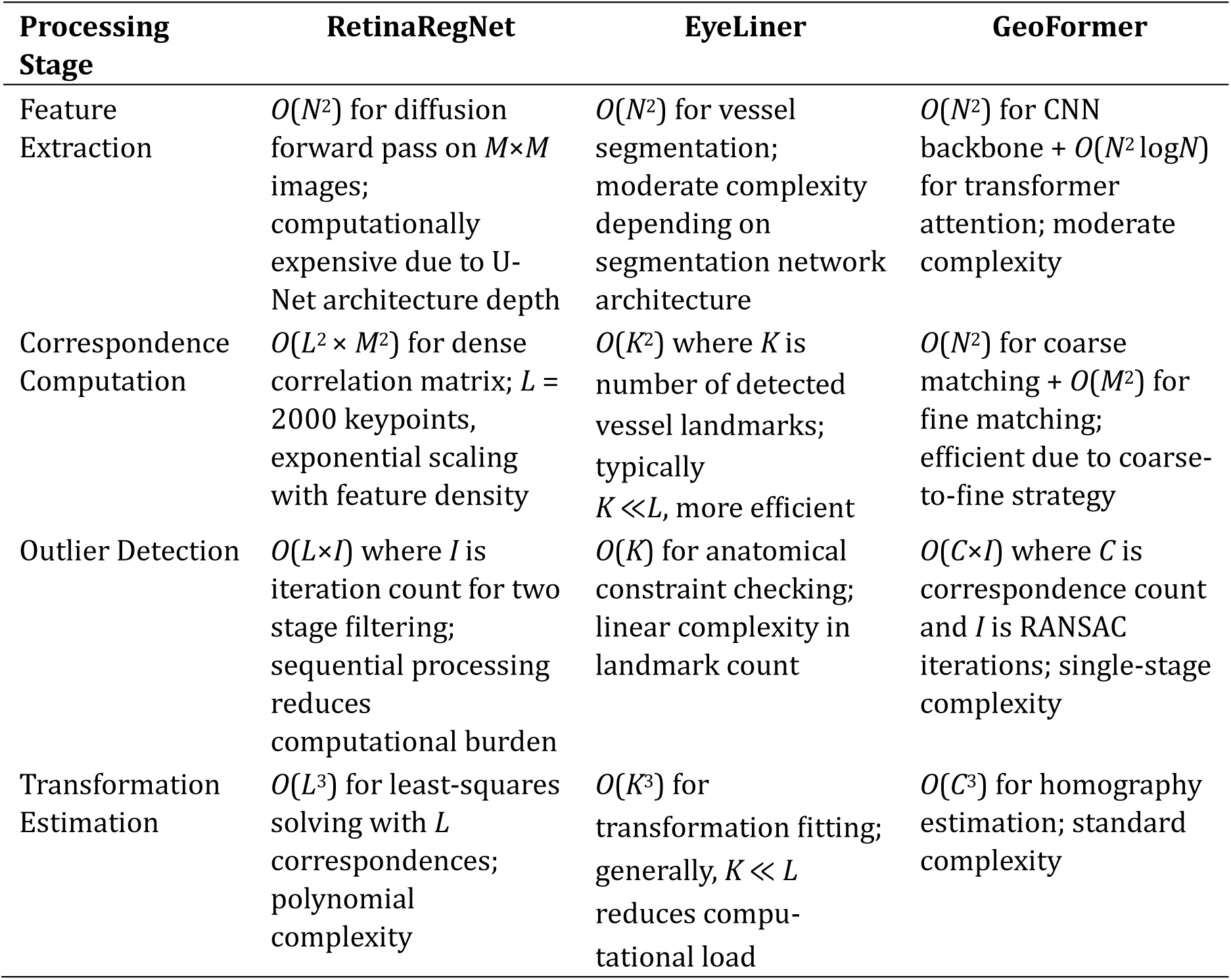
Algorithmic Complexity Analysis of Retinal Image Registration Methods.

Supplementary Table 1 presents the computational complexity analysis for each processing stage of the three retinal image registration methods, showing how algorithmic choices impact scalability and processing efficiency.

**Supplementary Table 2.** The four key stages in image registration, namely keypoint selection, keypoint matching, match outlier removal, and image warping, and the techniques applied by the three methods at each stage. The key methodological differences across RetinaRegNet, EyeLiner, and GeoFormer are summarised in Supplementary Table 2, outlining their respective strategies for keypoint selection, keypoint matching, outlier removal, and image warping.

|  | RetinaRegNet | EyeLiner | GeoFormer |
| --- | --- | --- | --- |
| Keypoint selection | SIFT + random sampling | SuperPoint + anatomy guided filtering | - |
| Keypoint matching | Dense 2D correlation on diffusion feature map | LightGlue | Attention mechanism |
| Match outlier removal | Inverse consistency + geometric transformation consistency | - | RANSAC + confidence thresholding + geometric locality constraint |
| Image warping | Homography global transformation + 3rd order polynomial local transformation | Affine global transformation + constrained local deformation | Homography transformation |

